# Intra-familial dynamics of mental distress during the Covid-19 lockdown

**DOI:** 10.1101/2024.12.04.24318462

**Authors:** Johanne H. Pettersen, Espen Eilertsen, Laura Hegemann, Laurie J. Hannigan, Ingunn Olea Lund, Pia M. Johannesen, Elizabeth C. Corfield, Eivind Ystrom, Ole A. Andreassen, Alexandra Havdahl, Ragnhild E. Brandlistuen, Helga Ask

## Abstract

**Importance:** Lockdowns and social restrictions imposed in response to the Covid-19 pandemic intensified the proximity and reciprocal exposure among members of nuclear families. It is unclear how variation in mental distress during this period is attributed to family members’ influence.

**Objective:** Using genetic data from family members, it is possible to disentangle parent-driven, child-driven, and partner-driven influences of mental distress. We separated between two types of influences; direct genetic–how an individual’s genotype influences their own mental distress, and indirect genetic–how an individual’s genotype influence the mental distress of family members.

**Design, Setting and Participants:** Based on a pre-registered analysis plan, we used longitudinal data from the Norwegian Mother, Father and Child Cohort Study, reported by adolescents (N=4,388), mothers (N=27,852) and fathers (N=25,953) during the first two months of the Covid-19 lockdown. Trio genome-wide complex trait analyses were used to separate direct and indirect genetic effects at each timepoint. Separate models also included adolescents’ non-pandemic responses (N=10,012). Trio polygenic score (PGS) design was used to investigate direct and indirect effects of specific liability factors, and whether the associations changed over days in lockdown, adding interaction terms (PGS×time).

**Exposure:** Family trios’ genotype and PGS for anxiety, depression, ADHD, neuroticism, and anorexia nervosa.

**Main Outcome:** Mental distress was measured using the five-item version of the Hopkins Symptom Checklist.

**Results:** We found evidence of family members influencing each other, with a pattern of greater proportions of variance explained later in lockdown. Indirect genetic effects were observed across family members; 10% of the variance in adolescent mental distress was mother-driven, 2-3% in mothers were partner-driven, and 5% in fathers were offspring-driven. Mothers’ genetic liability to depression and ADHD was positively associated with fathers’ mental distress. No interaction effects between PGS and time were found. Direct genetic effects contributed to 9-10% variance in mental distress across family members, partly explained by genetic variants associated with anxiety, depression, ADHD, and neuroticism.

**Conclusions and Relevance:** These findings highlight the importance of family dynamics, particularly during societal crises like the Covid-19 pandemic and emphasizes the value of including family members in mental health interventions.

**Key Points:** *Question:* How did mothers, fathers and adolescent offspring influence each other’s mental distress during the Covid-19 lockdown?

*Findings:* Up to 10% of the variance in mental distress was explained by family-related environmental factors (proxied by genetic variants in family members). Mother-driven effects were most important among adolescents, while father-driven effects explained variance among mothers, and child-driven effects explained variance in fathers. Fathers’ mental distress was also higher when mothers had higher genetic liability for ADHD and depression.

*Meaning:* The findings highlight the potential value of including family members in mental health interventions.

## Introduction

During the first weeks of the Covid-19 pandemic, many families were confined at home together. This abrupt environmental disruption, which for many also saw increased stress, uncertainty and mental distress^1,2^, provides a unique opportunity to examine the family dynamics of mental health.

Previous studies have suggested that family dynamics play a role in individuals’ mental distress (i.e., symptoms of anxiety and depression), even after adjusting for the genetic and environmental factors that are shared within families. The majority of this literature emphasizes what we can call *parent-driven effects*: how the role and behavior of parents shape the mental health of children, through for example parenting^3^ or maternal lifestyle during pregnancy^4,5^. Recently, such parental influences have been investigated using polygenic scores (PGS) related to specific traits (e.g., ADHD and depression). Parental PGS are included as exposures and offspring mental health as outcome in regression models. By adjusting such models for the child’s own PGS (the transmitted genetic variants), we can estimate of possible parent-driven effects on the child outcome. Such effects have been identified for common genetic variants associated with autism, ADHD, and educational attainment on child neurodevelopmental traits^6^, but with less evidence for other outcomes (e.g., on conduct problems^7^, and of the educational attainment PGS on children’s depressive, anxiety, or ADHD traits^8^).

Some literature also focuses on how romantic partners influence each other, through *partner-driven effects.* Although partner similarity across a range of traits is evident from the initial stages of a relationship (arising through processes like social homogamy and assortative mating), convergence over time has also been observed^9,10^. Partners likely influence each other’s mental health through their behavior and interactions. For example, relationship conflict and poor communication have been associated with higher maternal depression risk postpartum.^11^

Fewer studies investigate *child-driven effects* - how children influence the mental health of their parents. For instance, adoptions studies have shown that child emotional and behavioral problems are associated with mental health outcomes in adoptive parents^12,13^. In one study on the association between offspring sleep problems and maternal depression, child-driven effects appeared stronger than the mother-driven^14^. Child-driven effects have also been investigating using the trio-PGS design. For example, a recent study found evidence for both child- and partner-driven effects on maternal depression, through genetic liability to a general psychopathology factor, varying in strength across life stages.^15^

In addition to the search for specific traits or behaviors in one family member that influence mental health outcomes in another (trait-based approaches), variance-based approaches^16^ offer an alternative framework for investigating parent-, partner- and child-driven effects. In these approaches, specific variables related to traits or behaviors are not selected, but variance in the outcome of interest is partitioned based on its underlying source, estimated based on patterns of covariance across family members. Classical twin modelling is one example of a variance-based approach, disentangling variance explained by genetic, shared environmental and unique environmental influences. Twin studies have estimated the heritability of anxiety and depression at 30-50%,^17,18^ with shared environmental factors accounting for up to 30% of variance in childhood, though this influence typically diminishes in adulthood.^19^

Another recently developed variance-based approach is trio genome-wide complex trait analysis (Trio-GCTA),^20^ which uses data on genome-wide common genetic variants from mother, father, and offspring trios to separate direct and indirect genetic effects. Providing a broad perspective on the overall genetic contributions within families, the trio-GCTA offers a valuable initial step in understanding family dynamics, which can be complemented by subsequent trait-based approaches. Previous Trio-GCTA studies have estimated that partner- and child-driven effects account for between 0-14% variance in maternal depression across several timepoints,^21^ and that parent-driven effects explain 8-16% variance in conduct, inattention and hyperactivity^22^ in 8 year-old children. Mother-driven effects have also been shown to contribute 6-10% of variance in children’s early neurodevelopmental traits using this method^6^.

Most previous literature investigating parent-driven effects has relied on maternally reported child outcomes, potentially biasing the observed effects of mothers on children^6,22,23^. Therefore, investigating family dynamics using trio self-reported data is of importance. Also, few studies have examined family dynamics in families with older children (>8 years), an area of particular importance given the increased mental distress among adolescents. A recent systematic review described the Covid-19 pandemic as an intensifier of intergenerational risk of mental health problems among children^24^.

In this pre-registered study, we used longitudinal and cross-sectional data on mental distress reported by mother, father, and adolescent offspring trios during the first two months of the Covid-19 lockdown in Norway. Using both a variance-based (Trio-GCTA), and a trait-based (Trio-PGS) approach, we aimed to investigate 1) to what extent adolescents’, mothers’ and fathers’ mental distress was influenced by direct and indirect genetic effects 2) to what extent these effects could be explained by genetic variants associated with anxiety, depression, ADHD, neuroticism and anorexia and 3) whether the magnitude of direct and indirect effects changed over time in lockdown.

## Methods

### Study design and sample

The Norwegian Mother, Father, and Child Cohort Study (MoBa) is a population-based pregnancy cohort study conducted by the Norwegian Institute of Public Health.^25,26^ Participants were recruited from all over Norway from 1999-2008. The women consented to participation in 41% of the pregnancies. The cohort includes approximately 114,500 children, 95,200 mothers, and 75,200 fathers. The establishment of MoBa and initial data collection was based on a license from the Norwegian Data Protection Agency and approval from The Regional Committees for Medical and Health Research Ethics. The MoBa cohort is currently regulated by the Norwegian Health Registry Act. The current study was approved by The Regional Committees for Medical and Health Research Ethics (14140). Blood samples were obtained from both parents during pregnancy and from mothers and children (umbilical cord) at birth.^27^ Genotyping of MoBa, quality control, phasing, imputation, and post-imputation quality control has previously been described.^28^ MoBa data was linked to the Medical Birth Registry (MBRN), a national health registry containing information about all births in Norway.

Since March 2020, parents and 16-18-year-old adolescents in MoBa were invited to respond to biweekly Covid-19 questionnaires. Three of these questionnaires were sent out during the lockdown period in Norway between March and May 2020, including a measure of mental distress. In this study, we leveraged data on complete family trios of 15-18-year-old adolescents (N=4,388), mothers (N=27,852), and fathers (N=25,953) responding to at least one of these questionnaires. Mental distress among adolescents was also assessed in separate questionnaires sent out to 14-16-year-olds (from 2017-2023), resulting in a total sample of 13,484 adolescents. Trio-GCTA samples were restricted to unrelated trios (See flow chart in eFigure 1). This paper was preregistered in the open science framework (OSF): https://osf.io/2npgt. See eTable 1 for a complete list of deviations from the preregistration.

**Figure 1.**
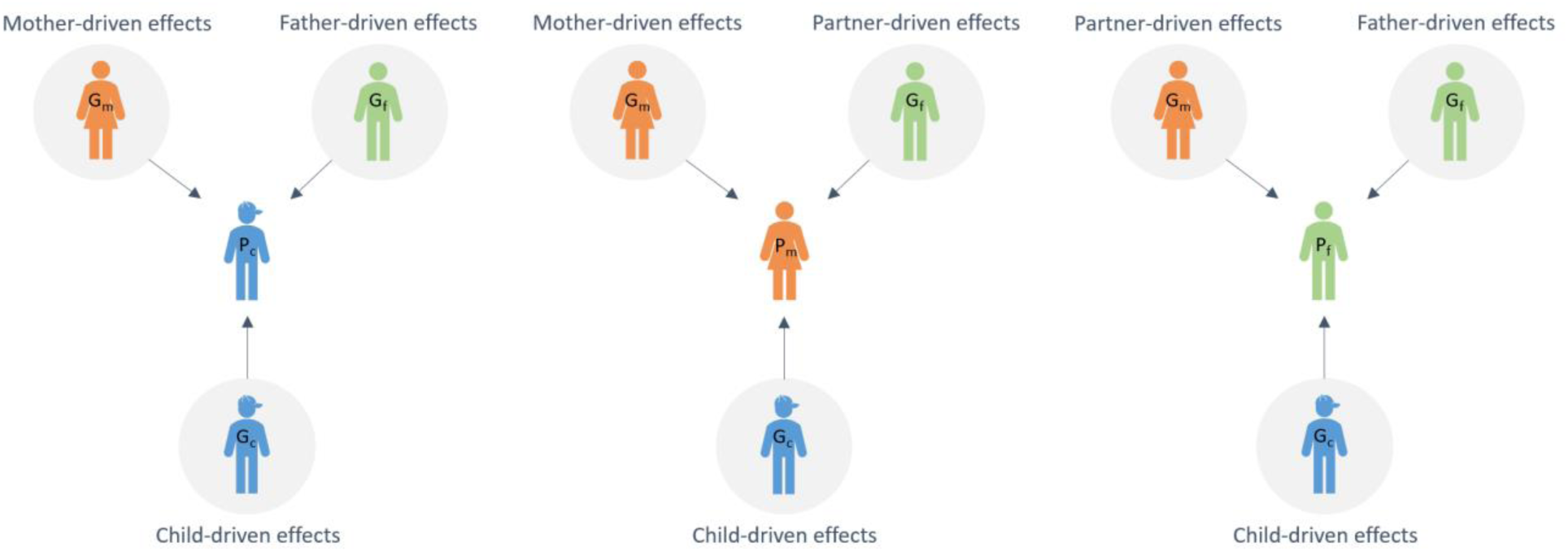
Illustrative model of mother-, father-, partner-, and child-driven effects on mental distress in family trios. **Note:** As the focal individual changes (mental distress in mother, father or adolescent), the interpretation of the effects changes. When the adolescent is the focal individual, the child-driven effects represent direct effects and mother- and father-driven effects are indirect genetic effects. When the mother/father is the focal individual the child- and partner-driven effects are the indirect effects and the mother/father-driven effects represent the direct genetic effects.

### Outcome

Mental distress was measured by the 5-item version of the Hopkin’s Symptom Checklist.^29^ Each item was scored on a scale of 1 (not bothered) to 4 (very bothered), based on experiences during the last two weeks. A mean SCL-5 score for each of the three timepoints during the lockdown period was estimated and multiplied by the total number of items. Additionally, an overall SCL-5 mean score was created for all adolescents (including 1-4 measurements). All SCL-5 scores were log-transformed to reduce skewness.

### Exposures

A PGS quantifies genetic liability to a particular condition by summing an individual’s risk alleles, each weighted by their effect size in a previous genome-wide association study (GWAS).^30^ PGS were created for anxiety,^31^ depression,^32^ neuroticism,^33^ ADHD^34^, and anorexia nervosa (AN)^35^ using LDpred2 software.^36^

Confounding variables included: adolescents’ sex assigned at birth (Male/Female), child age (in 2020, range 11-21-years), mothers age (30-63 years), and fathers age (31-75 years). Additionally, a continuous variable for time was created, representing the number of days between March 12^th^, 2020, and the completion of a given questionnaire. A variable on household status (trios living together) was created for sensitivity analyses. See eMethods for details on the variables and PGS.

### Statistical analyses

#### Variance-based approach

To disentangle the total direct and indirect genetic effects on mental distress in family trios, we used Trio-GCTA. This method uses a genomic relatedness matrix (GRM) based on common genome-wide SNPs from the individuals in the trio. The covariances between direct and indirect genetic effects can be interpreted as a gene-environment correlations.^20^ The Trio-GCTA method allows us to estimate unconfounded genetic effects of each family members on adolescent’s, mother’s and father’s mental distress.

For adolescents, due to limited power in the Covid-19 sample, one model was run including the overall SCL as outcome. For mothers and fathers, three separate models were run, one for each SCL measure during the Covid-19 lockdown. As the focal individual changes (adolescent, mother, or father), the interpretation of the model parameters also changes, as can be seen in the illustration in Figure 1.

Using the Julia programming language,^37^ and the VCModels.jl package^38^ we compared five models for each focal individual (adolescent, mother, father) and timepoint (see eTable 2 for variance components included in the different models). Each model included the fixed effects of child sex, genotype batch, and first 10 principal components.

The best-fitting model was selected based on the lowest Akaike’s Information Criteria (AIC)^39^ value. (See eMethods for further details).

#### Trait-based approach

Multilevel regression models, including the trio PGS as exposures and repeated measures of mental distress as outcome, were conducted in R^40^ using the lme4 package^41^. Separate models were run for each focal individual, and for each PGS (i.e., anxiety, depression, ADHD, neuroticism, and AN), 15 models in total. The indirect genetic effects were interpreted by the beta of the other family members PGS on mental distress, after adjusting for focal individual’s PGS (direct genetic effect).

To investigate whether the magnitude of genetic effects changed over time in lockdown, all models were rerun including interaction terms between each PGS and time.

All models were adjusted for time, age of the focal individual, and child’s sex. Random intercepts of the focal individuals ID were added to account for repeated measures. In the models with the adolescent’s outcome, we added a random intercept of maternal ID, to account for siblings. Full information maximum likelihood (FIML) was used to handle missing data. We used false discovery rate (FDR) to adjust for multiple testing for 5 tests (5 PGS). Supplementary trio-PGS analyses were run including all adolescents with available SCL-5 data. We also ran sensitivity trio-PGS analyses where we restricted the sample to family trios registered in the same household.

## Results

All participants reported highest level of mental distress at the first timepoint during lockdown (Madolescents=1.55,SD=0.60; Mmothers=1.46,SD=0.49; Mfathers=1.35,SD=0.44) and lowest at the third timepoint (Madolescents=1.52,SD=0.59; Mmothers=1.37,SD=0.46;Mfathers=1.31,SD=0.44).

Sociodemographic characteristics across samples can be found in Table 1.

**Table 1.** Sociodemographic characteristics Note: Mother’s and father’s education is measured as the highest completed level of education in 2020. SD = standard deviation; T1 = timepoint 1; T2 = timepoint 2; T3 = timepoint 3. Abbreviations: SCL-5 = Hopkins Symptom Checklist (5-item version).

| Sociodemographic characteristics |  | Adolescent trios |  | Mother trios |  |  | Father trios |  |  |
| --- | --- | --- | --- | --- | --- | --- | --- | --- | --- |
|  |  | Overall sample<br>(N = 13,484) | Lockdown sample<br>(N = 4,388) | T1<br>(N=25,575) | T2<br>(N=25,108) | T3<br>(N=23,591) | T1<br>(N =23,630) | T2<br>(N=22,285) | T3<br>(N=20,349) |
| **Observations** |  | 20,162 | 9,674 |  |  |  |  |  |  |
| **Child's sex** |  |  |  |  |  |  |  |  |  |
|  | Male | 6,211 (46.1%) | 1,878 (42.8%) | 12,971 (50.7%) | 12,732 (50.7%) | 11,955 (50.7%) | 12,046 (51.0%) | 11,374 (51.0%) | 10,344 (50.8%) |
|  | Female | 7,273 (53.9%) | 2,510 (57.2%) | 12,608 (49.3%) | 12,376 (49.3%) | 11,636 (49.3%) | 11,584 (40.0%) | 10,911 (49.0%) | 10,005 (49.2%) |
| **Mean (SD) age – Children** |  | 16.0 (1.1) | 16.9 (0.6) | 14.4 (2.0) | 14.4 (2.0) | 14.4 (2.0) | 14.3 (1.9) | 14.3 (2.0) | 14.3 (2.0) |
| **Mean (SD) age – Mothers** |  | 46.3 (4.4) | 47.1 (4.5) | 45.1 (4.8) | 45.2 (4.8) | 45.2 (4.8) | 45.0 (4.8) | 45.1 (4.7) | 45.2 (4.7) |
| **Mean (SD) age – Fathers** |  | 48.6 (5.1) | 49.4 (5.1) | 47.4 (5.5) | 47.5 (5.5) | 47.5 (5.5) | 47.4 (5.4) | 47.5 (5.4) | 47.6 (5.4) |
| **Mother's education** |  |  |  |  |  |  |  |  |  |
|  | Compulsory | 389 (2.9%) | 183 (4.2%) | 1,019 (4.0%) | 1,010 (4.0%) | 880 (3.7%) | 938 (4.0%) | 846 (3.8%) | 746 (3.7%) |
|  | Upper secondary | 2,712 (20.1%) | 1,144 (26.1%) | 5,779 (22.6%) | 5,689 (22.7%) | 5,195 (22.0%) | 5,181 (21.9%) | 4,787 (21.5%) | 4,299 (21.1%) |
|  | Bachelor's degree | 7,368 (54.6%) | 2,271 (51.2%) | 13,446 (52.6%) | 13,228 (52.7%) | 12,534 (53.1%) | 12,332 (52.2%) | 11,677 (52.4%) | 10,691 (52.5%) |
|  | Master's degree or Ph.D. | 3,008 (22.3%) | 788 (18.0%) | 5,314 (20.8%) | 5,165 (20.6%) | 4,967 (21.1%) | 5,158 (21.8%) | 4,961 (22.3%) | 4,605 (22.6%) |
| **Father's education** |  |  |  |  |  |  |  |  |  |
|  | Compulsory | 767 (5.7%) | 289 (6.6%) | 1,712 (6.7%) | 1,674 (6.7%) | 1,518 (6.4%) | 1,358 (5.7%) | 1,234 (5.5%) | 1,078 (5.3%) |
|  | Upper secondary | 5,123 (20.1%) | 1,891 (43.1%) | 10,272 (40.2%) | 10,118 (40.3%) | 9,352 (39.6%) | 8,951 (37.9%) | 8,259 (37.1%) | 7,472 (36.7%) |
|  | Bachelor's degree | 4,598 (34.1%) | 1,383 (31.5%) | 8,385 (32.8%) | 8,263 (32.9%) | 7,861 (33.3%) | 8,160 (34.5%) | 7,801 (35.0%) | 7,180 (35.3%) |
|  | Master's degree or Ph.D. | 2,980 (22.1%) | 823 (18.8%) | 5,157 (20.2%) | 5,005 (19.9%) | 4,816 (20.4%) | 5,132 (21.7%) | 4,968 (22.3%) | 4,598 (22.6%) |
| **Mean (SD) scl-5 – Adolescents** |  | 1.56 (0.62) | 1.54 (0.59) |  |  |  |  |  |  |
| **Mean (SD) scl-5 – Mothers** |  |  |  | 1.46 (0.49) | 1.42 (0.47) | 1.37 (0.46) |  |  |  |
| **Mean (SD) scl-5 – Fathers** |  |  |  |  |  |  | 1.35 (0.44) | 1.34 (0.45) | 1.31 (0.44) |
| **SCL-5 cut off (2.0)** |  |  |  |  |  |  |  |  |  |
|  | No | 15,346 (76.1%) | 7,411 (76.6%) | 21,106 (82.5%) | 21,003 (83.7%) | 20,139 (85.4%) | 20,674 (87.5%) | 19,295 (86.6%) | 17,720 (87.1%) |
|  | Yes | 4,818 (23.9%) | 2,265 (23.4%) | 4,469 (17.5%) | 4,105 (16.3%) | 3,452 (14.6%) | 2,956 (12.5%) | 2,990 (13.4%) | 2,629 (12.9%) |

### Trio-GCTA

#### Parental models

For mothers, the best-fitting model at the first timepoint included only direct genetic effects, explaining 9.9% variance in mental distress. For the second and third timepoints, models including direct and father indirect effects revealed the best fit, with father indirect effects contributing 2.9% and 2.6% and direct effects explaining 9.5% and 9.3% of the variance, respectively (eTable 3).

For fathers, the best-fitting model at timepoint 1-2 included only direct effects, explaining 10.4% and 10.2% variance, respectively. At timepoint 3, the optimal model included child indirect and direct effects, explaining 4.6% and 10.1%, respectively, with a negative covariance between these (correlation of -0.49). Figure 2 shows variance explained in each model.

**Figure 2.**
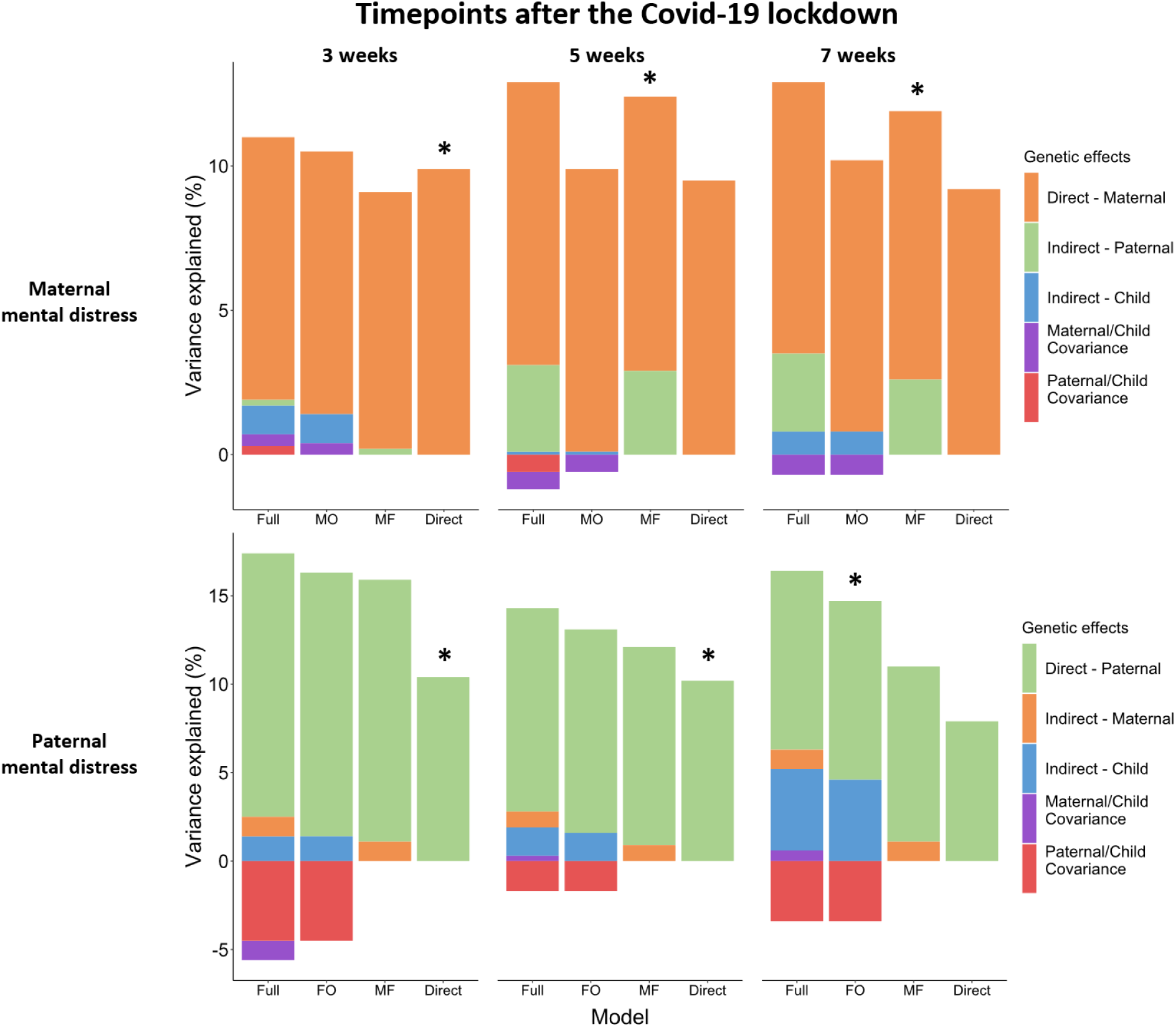
Trio-GCTA model run on mothers’ and fathers’ mental distress measured at three timepoints during the Covid-19 lockdown. **Note:** The trio-GCTA model on maternal and paternal mental distress. Variance component estimates are presented for each model tested: full model with all parameters included (Full); maternal/paternal direct and child indirect effects (MO/FO); and direct effect model including maternal/paternal genetic effect (Direct). The best-fitting models based on the lowest AIC is indicated by *. Covariance between maternal and paternal effects is estimated in the model but not shown in the figure as we do not expect it to contribute to the variance in maternal/paternal mental distress.

#### Adolescent models

The model estimating direct and maternal indirect genetic effects was best-fitting for adolescent mental distress (eTable 4), explaining 8.8 % and 10.4%, respectively. There was a negative covariance between maternal indirect and direct genetic effects (correlation of -0.13). Figure 3 shows variance explained in each model.

**Figure 3.**
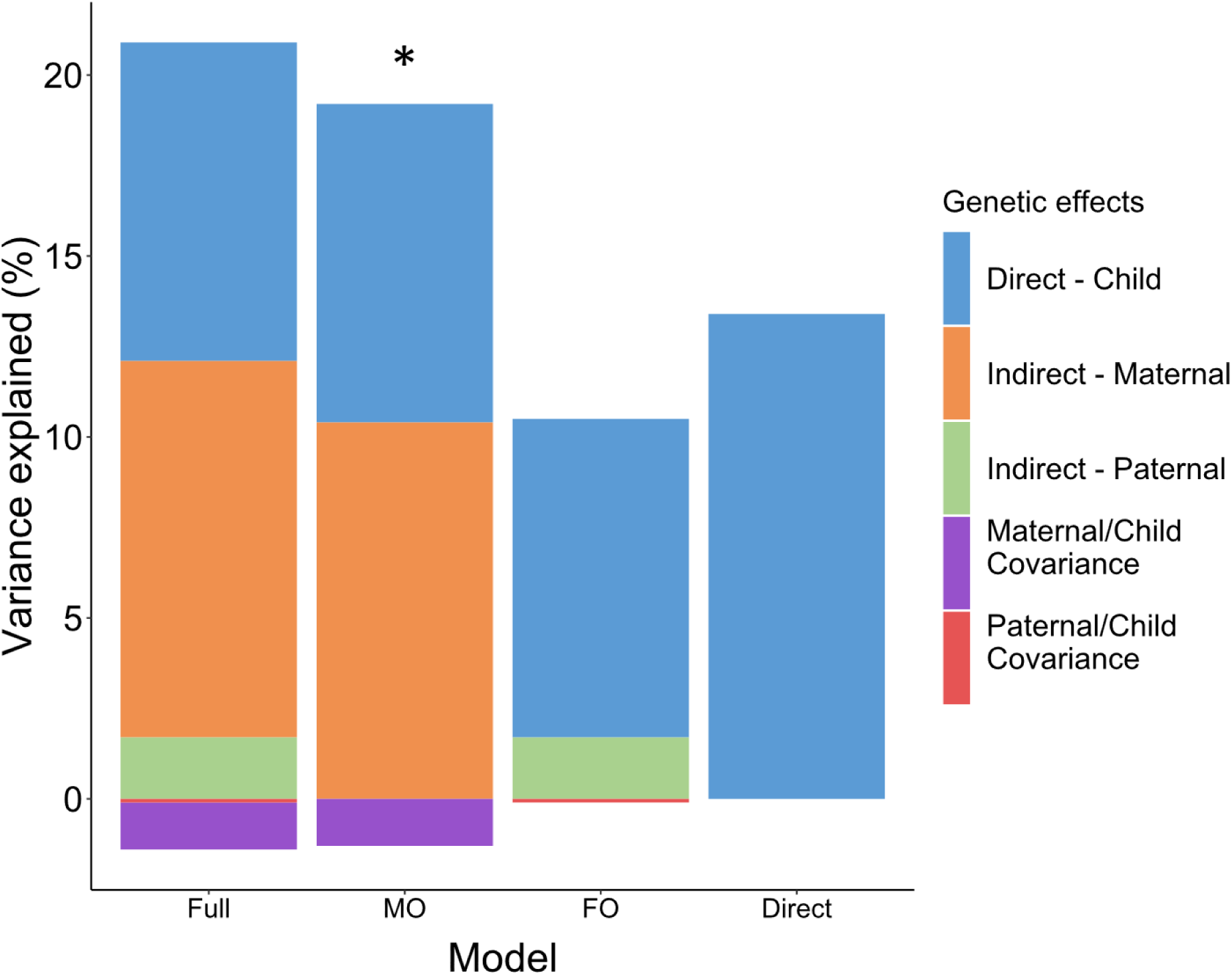
Trio-GCTA model run on adolescents’ mental distress. **Note:** Variance component estimates are estimated for each model tested: full model with all parameters included (Full model); child direct and mother indirect effects (MO); child direct and father indirect effects (FO); direct effects model only including the child’s genetic effect (Direct). The best-fitting models based on the lowest AIC are indicated by *.

### PGS analyses

The PGS analyses on father’s mental distress revealed indirect genetic effects of mother’s depressionPGS (β=0.02, 95%CI[0.01,0.04]) and ADHDPGS (β=0.03[0.02,0.04]). There were no significant indirect genetic effects on adolescent or maternal mental distress.

Direct genetic effects were observed across all models for depressionPGS (βadolescent=0.08[0.05,0.12];βmother=0.11[0.09,0.12];βfather=0.10[0.09,0.12]), and neuroticismPGS (βadolescent=0.07[0.03,0.11];βmother=0.11[0.09,0.12];βfather=0.10[0.08,0.11]). We also found direct genetic effects of anxietyPGS (βmother=0.05[0.03,0.06];βfather=0.05[0.04,0.06]), ADHDPGS (βmother=0.03[0.01,0.04]; βfather=0.04[0.02,0.05]) and ANPGS (βmother=0.03[0.02,0.04]; βfather=0.03[0.01,0.04]) on parental mental distress.

We found no significant interaction effects between PGS and time in lockdown. Standardized estimates can be found in eTable 5 and Figure 4.

**Figure 4.**
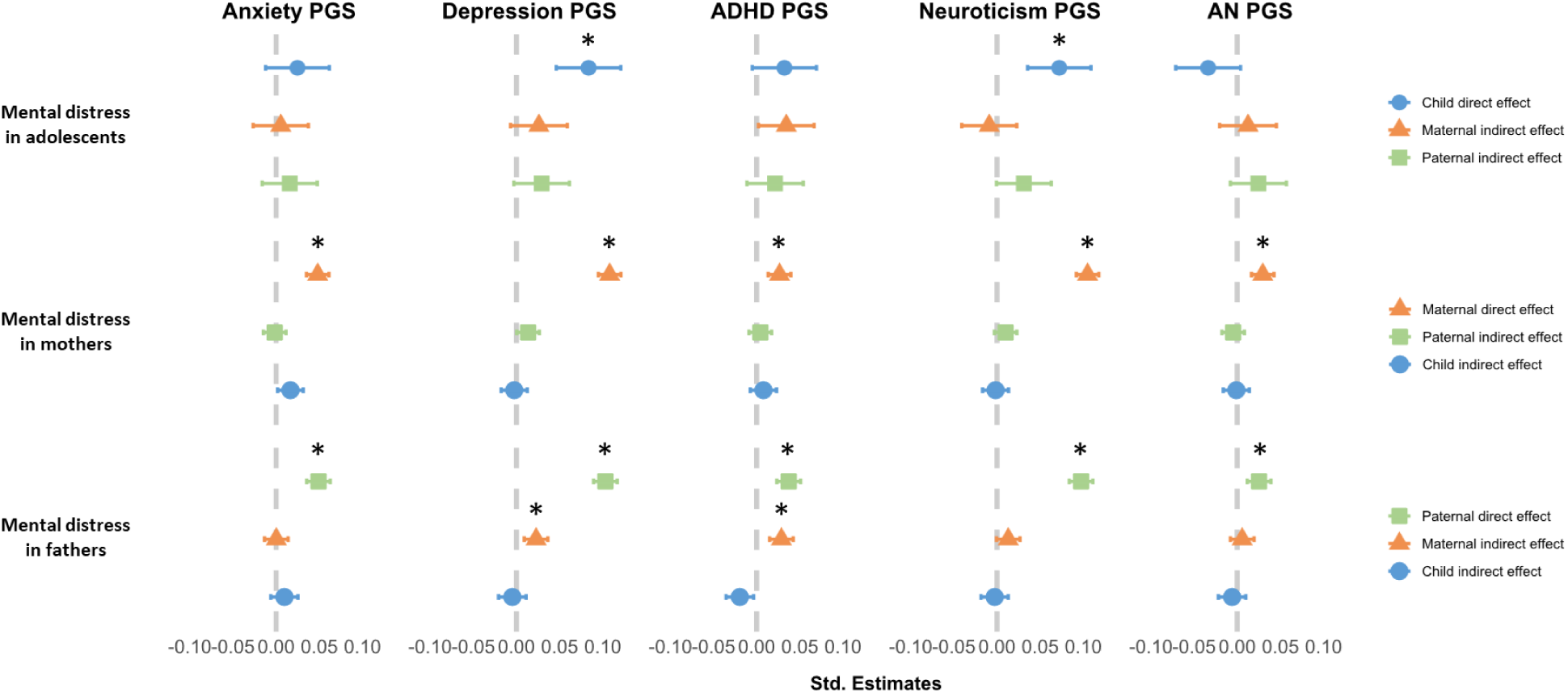
Standardized beta estimates for PGS on adolescents, mothers, and fathers’ mental distress. **Note:** Blue circles show adolescent genetic effects, maternal genetic effects are indicated by orange triangles, and paternal by green squares. * Indicates significant p-values after FDR correction.

Supplementary trio-PGS analyses on adolescent overall mental distress (not restricted to pandemic measures), revealed only direct genetic influences (eTable 6 and eFigure 2). Sensitivity analyses including only family members registered in the same household revealed similar results (see eTable 7).

## Discussion

In this pre-registered study, we used two complementary approaches to examine within-family processes and mental distress in parents and adolescent offspring during the first Covid-19 lockdown in 2020 in Norway. First, we quantified the total variance in mental distress explained by indirect and direct genetic influences. Next, we investigated to what extent these associations were captured by genetic liability to anxiety, depression, ADHD, neuroticism, and AN. Last, we investigated whether these effects changed over time. We found evidence for both indirect and direct genetic influences and, while formal comparisons detected no interactions with time spent in lockdown, an increasing proportion of variance was accounted for by IGEs at later waves.

### Mothers are important for adolescent mental distress

In line with previous results based on younger children,^23,42^ more than 10% of the variation in adolescent mental distress (measured before and during the pandemic) was linked to mothers’ genes. Previous research has indicated that indirect genetic effects may decline as children mature; for example, a decrease in the influence of shared environments with age has been repeatedly observed in twin studies^19^. The mother-driven influences observed in younger children has been suggested to be partly explained by a maternal rating bias, as the mother often report on the mental health outcomes of younger children. Our finding of mother-driven (and lack of father-driven) influences on adolescent *self-reported* mental distress is therefore particularly novel.

Mother-driven effects are supposedly mediated through the mother’s behavior or the family environment she provides. For instance, a genetic predisposition to neuroticism could affect parenting style, which in turn could impact child mental health. A study on 8-year-old children (in MoBa), suggested that maternal anxiety and depression symptoms partially mediated the indirect maternal effects.^23^ In our trio-PGS analyses, however, we did not find support for indirect maternal effects for any of the included PGS. On the other hand, the correlation between the child genetic effect and maternal indirect effect (gene-“environment” correlation) was close to zero in the trio-GCTA model. Based on this lack of correlation we would not expect the same PGS to be important for both indirect and direct genetic effects. Hence, the mother-driven associations (observed in the trio-GCTA results) could be explained by traits and conditions not included in our PGS analyses. For example, reflecting environmental influences in utero and during childhood.

### Child- and partner-driven effects on parental mental distress

We found no evidence for indirect genetic effects in parental mental distress at the first timepoint. However, in the following two bi-weekly data collections, father-driven effects accounted for 2- 3% of the variance in mothers and child-driven effects explained 5% of variance in fathers’ mental distress at the third timepoint.

Our results build on and extend previous research that report between 0-14% variance in maternal depression to be explained by indirect genetic effects (from both child and partner), when measured between the child was 6 months and 8 years old^21^. Uniquely, our study separates the genetic contributions of children and partners.

Our trait-based approach did not support that the included PGS explained the observed father- driven and child-driven effects. However, we did find evidence for an indirect genetic effect of maternal depression and ADHD genetic liability on paternal mental distress. Higher genetic liability to depression or ADHD in mothers was associated with elevated levels of mental distress in fathers. In the full (not the best-fitting) Trio-GCTA model for fathers’ mental distress, indirect maternal effects explain 1% of the variance, supporting that such influences could exist. The PGS generally explain very little variance in the outcome. Therefore, the indirect effects from the PGS would also be too small to be detected in the larger picture we get from the trio-GCTA models.

Trio-GCTA and trio-PGS, while complementary, might yield inconsistent results as they operate at different scales.

### Direct genetic effects

As expected by previous studies^21,23^, direct genetic effects explained 9-10% variation in mental distress across models. These numbers are comparable to previous results from the same cohort. For example, direct effects explained 5-19% of variance in anxiety and depression symptoms at age 8^23^ and 5-14% in maternal depression symptoms^21^. Trait-based analyses suggested that the direct effect could partly be explained by genetic liability to the included traits, particularly depression and neuroticism. Consistent with previous studies^21^, our analyses revealed a negative covariance between the direct and indirect effects on adolescent mental distress, suggesting that the same genetic variants in mothers and their children may have opposing influences on mental distress, highlighting a possible gene-environment correlation.^20^

### Time in lockdown

Evidence from our study as to whether or not time in lockdown changed the intra-familial dynamics of mental distress was mixed. While we observed a pattern of increasing proportion variance explained on parental mental distress in the trio-GCTA models across time in lockdown, we did not find that the direct and indirect genetic effects changed as a function of time in lockdown in the trio-PGS models. However, most of the family-driven effects were not captured by the included PGS. The pattern of increase in indirect genetic effects seen across time in the trio-GCTA models might reflect other processes than those captured by the PGS.

It is uncertain whether the pandemic lockdown shaped the findings on adolescent mental distress. Due to power limitations, we included data on adolescents reporting mental distress outside of the pandemic. However, the pandemic setting could still drive the observed indirect maternal effect. Further research is needed to examine all associations independently of the pandemic lockdown.

### Limitations

Our study has some limitations that are relevant to consider when interpreting our results. First, in the trio-GCTA model comparisons, model fit revealed small differences, and the statistical support for the selected best-fitting models might be limited. Second, the MoBa sample consists of higher educated and healthier families^43^, and limiting our analyses to complete trios have likely resulted in a highly selected sample. Third, factors like assortative mating, residual population stratification, and sibling effects are likely to be captured in our estimates of indirect genetic effects, limiting the extent to which they can be interpreted as evidence of exposure-based environmental effects^22^. Finally, our data were based on a quality control restricted to participants of European ancestry, limiting our results’ generalizability.

## Conclusion

Our findings provide evidence for intra-familial processes in shaping mental distress. Mother-driven effects explained more variance than direct genetic effects in adolescent mental distress. Father-driven effects contributed to maternal mental distress, and mental distress in fathers was influenced by all family members. These findings highlight the complex interplay of genetic and environmental factors in shaping mental health within nuclear families, particularly during periods of prolonged environmental stressors such as the Covid-19 lockdown. Understanding these dynamics is important in designing mental health interventions.

## Article Information

### Author Contributions

Ms Pettersen had full access to the data in the study and takes responsibility for the integrity of the data and the accuracy of the data analysis.

*Concept and design:* Pettersen, Hannigan, Eilertsen, Ask *Acquisition, analysis, or interpretation of data:* All authors *Drafting of the manuscript:* Pettersen

*Critical review of the manuscript for important intellectual content:* All authors

*Statistical analysis:* Eilertsen, Pettersen

*Obtained funding:* Brandlistuen, Ask

*Administrative, technical, or material support:* Lund, Ask

*Supervision:* Brandlistuen, Eilertsen, Ask

### Data Sharing Statement

Analysis code is publicly available on GitHub: https://github.com/psychgen/Family-trios-Covid19.

## Supporting information

eFigure

## Data Availability

Data from MoBa used in this study are managed by the national health register holders in Norway (Norwegian Institute of public health) and can be made available to researchers, provided approval from the Regional Committees for Medical and Health Research Ethics (REC), compliance with the EU General Data Protection. Regulation (GDPR) and approval from the data owners. The consent given by the participants does not open for storage of data on an individual level in repositories or journals. Researchers who want access to data sets for replication should apply through helsedata.no. Access to datasets requires approval from REC in Norway and an agreement with MoBa.

## References

1. Racine N, McArthur BA, Cooke JE, Eirich R, Zhu J, Madigan S. Global prevalence of depressive and anxiety symptoms in children and adolescents during COVID-19: a meta-analysis. JAMA pediatrics. 2021;175(11):1142–1150.

2. Madigan S, Racine N, Vaillancourt T, et al. Changes in Depression and Anxiety Among Children and Adolescents From Before to During the COVID-19 Pandemic: A Systematic Review and Meta-analysis. JAMA pediatrics. 2023;

3. Gorostiaga A, Aliri J, Balluerka N, Lameirinhas J. Parenting styles and internalizing symptoms in adolescence: a systematic literature review. International journal of environmental research and public health. 2019;16(17):3192.

4. Gustavson K, Ystrom E, Stoltenberg C, et al. Smoking in pregnancy and child ADHD. Pediatrics. 2017;139(2)

5. Gustavson K, Ystrom E, Ask H, et al. Acetaminophen use during pregnancy and offspring attention deficit hyperactivity disorder–a longitudinal sibling control study. JCPP advances. 2021;1(2):e12020.

6. Hegemann L, Eilertsen E, Pettersen JH, et al. Direct and Indirect Genetic Effects on Early Neurodevelopmental Traits. medRxiv. 2024:2024.01. 24.24301734.

7. Frach L, Barkhuizen W, Allegrini AG, et al. Examining intergenerational risk factors for conduct problems using polygenic scores in the Norwegian Mother, Father and Child Cohort Study. Mol Psychiatry. 2024;29(4):951–961.

8. Hughes AM, Torvik FA, van Bergen E, et al. Parental education and children’s depression, anxiety, and ADHD traits, a within-family study in MoBa. npj Science of Learning. 2024;9(1):46.

9. Ask H, Rognmo K, Torvik FA, Røysamb E, Tambs K. Non-random mating and convergence over time for alcohol consumption, smoking, and exercise: the Nord-Trøndelag Health Study. Behav Genet. 2012;42:354–365.

10. Ask H, Idstad M, Engdahl B, Tambs K. Non-random mating and convergence over time for mental health, life satisfaction, and personality: The Nord-Trøndelag Health Study. Behav Genet. 2013;43:108–119.

11. Pilkington PD, Milne LC, Cairns KE, Lewis J, Whelan TA. Modifiable partner factors associated with perinatal depression and anxiety: A systematic review and meta-analysis. J Affect Disord. 2015;

12. Ahmadzadeh YI, Eley TC, Leve LD, et al. Anxiety in the family: A genetically informed analysis of transactional associations between mother, father and child anxiety symptoms. Journal of Child Psychology and Psychiatry. 2019;60(12):1269–1277.

13. McAdams TA, Rijsdijk FV, Neiderhiser JM, et al. The relationship between parental depressive symptoms and offspring psychopathology: evidence from a children-of-twins study and an adoption study. Psychol Med. 2015;45(12):2583–94. doi:10.1017/S0033291715000501

14. Ystrom H, Nilsen W, Hysing M, Sivertsen B, Ystrom E. Sleep problems in preschoolers and maternal depressive symptoms: An evaluation of mother-and child-driven effects. Dev Psychol. 2017;53(12):2261.

15. Ayorech Z, Cheesman R, Eilertsen EM, et al. Maternal depression and the polygenic p factor: A family perspective on direct and indirect effects. Journal of Affective Disorders. 2023;332:159–167.

16. Bijma P. The quantitative genetics of indirect genetic effects: a selective review of modelling issues. Heredity (Edinb). Jan 2014;112(1):61–9. doi:10.1038/hdy.2013.15

17. Kendall K, Van Assche E, Andlauer T, et al. The genetic basis of major depression. Psychol Med. 2021;51(13):2217–2230.

18. Smoller JW. The genetics of stress-related disorders: PTSD, depression, and anxiety disorders. Neuropsychopharmacology. 2016;41(1):297–319.

19. Cheesman R, Rayner C, Eley T. The genetic basis of child and adolescent anxiety. Pediatric anxiety disorders. Elsevier; 2019:17–46.

20. Eilertsen EM, Jami ES, McAdams TA, et al. Direct and Indirect Effects of Maternal, Paternal, and Offspring Genotypes: Trio-GCTA. Behav Genet. 2021/03/01 2021;51(2):154-161. doi:10.1007/s10519-020-10036-6

21. Bjørndal LD, Eilertsen EM, Ayorech Z, et al. Disentangling direct and indirect genetic effects from partners and offspring on maternal depression using trio-GCTA. Nature Mental Health. 2024:1–9.

22. Eilertsen EM, Cheesman R, Ayorech Z, et al. On the importance of parenting in externalizing disorders: an evaluation of indirect genetic effects in families. Journal of child psychology and psychiatry. 2022;63(10):1186–1195.

23. Cheesman R, Eilertsen EM, Ahmadzadeh YI, et al. How important are parents in the development of child anxiety and depression? A genomic analysis of parent-offspring trios in the Norwegian Mother Father and Child Cohort Study (MoBa). BMC Med. Oct 27 2020;18(1):284. doi:10.1186/s12916-020-01760-1

24. Stracke M, Heinzl M, Müller AD, et al. Mental health is a family affair—systematic review and meta-analysis on the associations between mental health problems in parents and children during the COVID-19 pandemic. International Journal of Environmental Research and Public Health. 2023;20(5):4485.

25. Magnus P, Irgens LM, Haug K, Nystad W, Skjærven R, Stoltenberg C. Cohort profile: The Norwegian Mother and Child Cohort Study (MoBa). Int J Epidemiol. 2006;35(5):1146–1150. doi:10.1093/ije/dyl170

26. Magnus P, Birke C, Vejrup K, et al. Cohort Profile Update: The Norwegian Mother and Child Cohort Study (MoBa). Int J Epidemiol. 2016;45(2):382–388. doi:10.1093/ije/dyw029

27. Paltiel L, Anita H, Skjerden T, et al. The biobank of the Norwegian Mother and Child Cohort Study–present status. Norsk epidemiologi. 2014;24(1-2)

28. Corfield EC, Frei O, Shadrin AA, et al. The Norwegian Mother, Father, and Child cohort study (MoBa) genotyping data resource: MoBaPsychGen pipeline v.1. Cold Spring Harbor Laboratory; 2022.

29. Tambs K, Moum T. How well can a few questionnaire items indicate anxiety and depression? Acta Psychiatr Scand. 1993;87(5):364–367.

30. Choi SW, Mak TS, O’Reilly PF. Tutorial: a guide to performing polygenic risk score analyses. Nat Protoc. Sep 2020;15(9):2759–2772. doi:10.1038/s41596-020-0353-1

31. Purves KL, Coleman JR, Meier SM, et al. A major role for common genetic variation in anxiety disorders. Mol Psychiatry. 2020;25(12):3292–3303.

32. Adams MJ. Trans-ancestry genome-wide study of depression identifies 697 associations implicating cell-types and pharmacotherapies (in press). 2024.

33. Nagel M, Jansen PR, Stringer S, et al. Meta-analysis of genome-wide association studies for neuroticism in 449,484 individuals identifies novel genetic loci and pathways. Nat Genet. 2018;50(7):920–927.

34. Demontis D, Walters GB, Athanasiadis G, et al. Genome-wide analyses of ADHD identify 27 risk loci, refine the genetic architecture and implicate several cognitive domains. Nat Genet. 2023;55(2):198–208.

35. Watson HJ, Yilmaz Z, Thornton LM, et al. Genome-wide association study identifies eight risk loci and implicates metabo-psychiatric origins for anorexia nervosa. Nat Genet. 2019;51(8):1207–1214. doi:10.1038/s41588-019-0439-2

36. Privé F, Arbel J, Vilhjálmsson BJ. LDpred2: better, faster, stronger. Bioinformatics. 2020;36(22- 23):5424–5431.

37. Bezanson J, Edelman A, Karpinski S, Shah VB. Julia: A fresh approach to numerical computing. SIAM review. 2017;59(1):65–98.

38. Eilertsen EM. VCModels (1.0.0) [Julia]. https://github.com/espenmei/VCModels.jl

39. Akaike H. Factor analysis and AIC. Psychometrika. 1987;52:317–332.

40. *R: A Language and Environment for Statistical Computing*. R Foundation for Statistical Computing; 2021. https://www.R-project.org/

41. Bates D, Maechler M, Bolker B, et al. Package ‘lme4’. URL http://lme4 *r-forge r-project org*. 2009;

42. Ask H, Eilertsen EM, Gjerde LC, et al. Intergenerational transmission of parental neuroticism to emotional problems in 8-year-old children: Genetic and environmental influences. JCPP advances. 2021;1(4):e12054.

43. Nilsen RM, Vollset SE, Gjessing HK, et al. Self-selection and bias in a large prospective pregnancy cohort in Norway. Paediatr Perinat Epidemiol. 2009;23(6):597–608.

