## Supplementary material for "Intra-familial dynamics of mental distress during the Covid-19 lockdown": eFigure

**eTable 1:** Deviations from the preregistered analysis plan

| # | Details | | Original wording | Deviation Description | Reader Impact |
|---|---|---|---|---|---|
| 1 | Type | Research questions | Indirect genetic influences on mental distress in mother-father-offspring trios during the Covid-19 lockdown | We recognized the limited number of studies using self-reported measures from adolescents. To address this gap in the literature, we decided to also include direct genetic effects as our interest scope. | Including results of direct genetic effects provides a more comprehensive understanding of mental distress within and among family members. Although the analytical approach remained unchanged, this adjustment allows for a broader interpretation of the results. |
| | Reason | New knowledge | | | |
| | Timing | After data access | | | |
| 2 | Type | Analysis | The trio-GCTA model requires genetic data on each member of the family trio (mother, father, and offspring) and SCL-5 on each focal individual (family member of interest). Sample sizes in analyses for H1a-H1b will be limited to trios with SCL-5 measures for offspring, H1c-H1d will be limited to trios with SCL-5 measures for mothers, and H1e-H1f will be limited to trios with SCL-5 measures for fathers. | We initially planned to only use data collected during the first months of the Covid-19 lockdown and run a trio-GCTA model on adolescents. As the trio-GCTA model requires a larger sample size, the adolescents' sample was underpowered. We included adolescents responding prior to the pandemic to enhance power to run the Trio-GCTA. | Although we were not able to run the trio-GCTA model on adolescents reports only during the Covid-19 lockdown, this change broadens the scope of the analysis, providing a more nuanced understanding of mental distress in adolescents by including pre-pandemic data. |
| | Reason | Preregistered plan not possible or inappropriate because of data | | | |
| | Timing | After data access | | | |
| 3 | Type | Analysis | For the adolescents we will run a longitudinal trio-GCTA model to investigate H2, which will include data from the three SCL-5 timepoints. | We required assistance from a co-author to help develop a longitudinal trio-GCTA. We realized that this was not possible within the scope of this project, and therefore decided to run three trio-GCTA models (one for each timepoint) to capture change over time. | This adjustment ensures that readers gain valuable insight into the progression of direct and indirect genetic effects across timepoints, even without a fully integrated longitudinal model. |
| | Reason | Preregistered plan not possible or inappropriate because of data | | | |
| | Timing | After data access | | | |

| | | | | | |
|---|---|---|---|---|---|
| 4 | Type | Analysis | We will restrict our sample to family members living in the same household. | We intended to restrict our sample to family members living in the same household. However, the variable we received from Statistics Norway (SSB) was not optimal for this purpose and contained much missing data. Due to the need for large sample sizes, we decided to run the trio-GCTA models without this restriction. To account for potential household effects, we included sensitivity analyses in the trio-PGS models by restricting to trios registered as living in the same household. | While the main trio-GCTA analyses include all trios, the additional sensitivity analyses ensure that the readers can assess whether household status might have influenced the results. |
| | Reason | Pre-registered plan not possible or inappropriate because of data | | | |
| | Timing | After data access | | | |
| 5 | Type | Analysis | Exploratory analyses including mediator variables such as psychiatric disorders and primary health care visits (information from Norwegian Patient Registry (NPR) and The Norwegian Control and Payment of Health Reimbursement register (KUHR)) will be run for all the hypotheses with significant findings. | Due to the extensive number of analyses already included in this paper, we decided not to incorporate these exploratory analyses at this stage. | By choosing to focus on the primary analyses, and supplementary/sensitivity analyses already included the readers will have a clearer, more focused interpretation of the main results. |
| | Reason | New knowledge | | | |
| | Timing | After data analyses | | | |
| 6 | Type | Analysis | All models will be adjusted for child's sex and parental age. Children's age will be included as a covariate in models with parental mental distress as the outcome. | After reviewing the existing literature, we realized that the common practice is to adjust trio-GCTA models for primarily for child sex, batch, and principal components. Given the complexity of these models, we decided to avoid over-adjustment. Regarding the trio-PGS models, | We do not believe this deviation would impact the results as we have little evidence for believing parents age would influence the strength of the indirect effects. |
| | Reason | Typo/oversight | | | |
| | Timing | After data access | | | |

| # | Details | | Original wording | Deviation Description | Reader Impact |
|---|---|---|---|---|---|
| | | | | these were adjusted for focal individuals age, time, and child sex. | |
| 7 | | Analyses | PRSice2 will be used to create polygenic scores (PGS) for each family member based on summary statistics from genome-wide association studies (GWAS). | LDpred2 has emerged as a more predictive and robust method of calculating PGS on complex traits. We therefore chose to select this method instead of PRsice2 to enhance predictive power. | This change benefits the readers by improving the accuracy and robustness of the findings related to genetic liability. |
| | | New knowledge | | | |
| | | After data access | | | |
| **Unregistered steps** | | | | | |
| **#** | **Details** | | **Original wording** | **Deviation Description** | **Reader Impact** |
| 1 | Type | Variables | We did not specify including polygenic score of anorexia nervosa in the preregistration. | Although we did not specify adding anorexia nervosa as one of the PGS traits, we believe that this addition is relevant as there has been an increase in eating disorders during the Covid-19 pandemic highlighting the relevance of including this PGS. | This deviation adds to the depth to the analysis by considering a genetic component that may influence the traits under study. Readers will gain a more nuanced understanding of how genetic liability to anorexia nervosa might contribute to mental distress. |
| | Reason | New knowledge | | | |
| | Timing | After data access | | | |
| | Reason | New knowledge | | | |
| | Timing | After data access | | | |

**Note:** Following the Preregistration Deviation Table Template[1]

**eFigure 1:** Flow chart of study samples

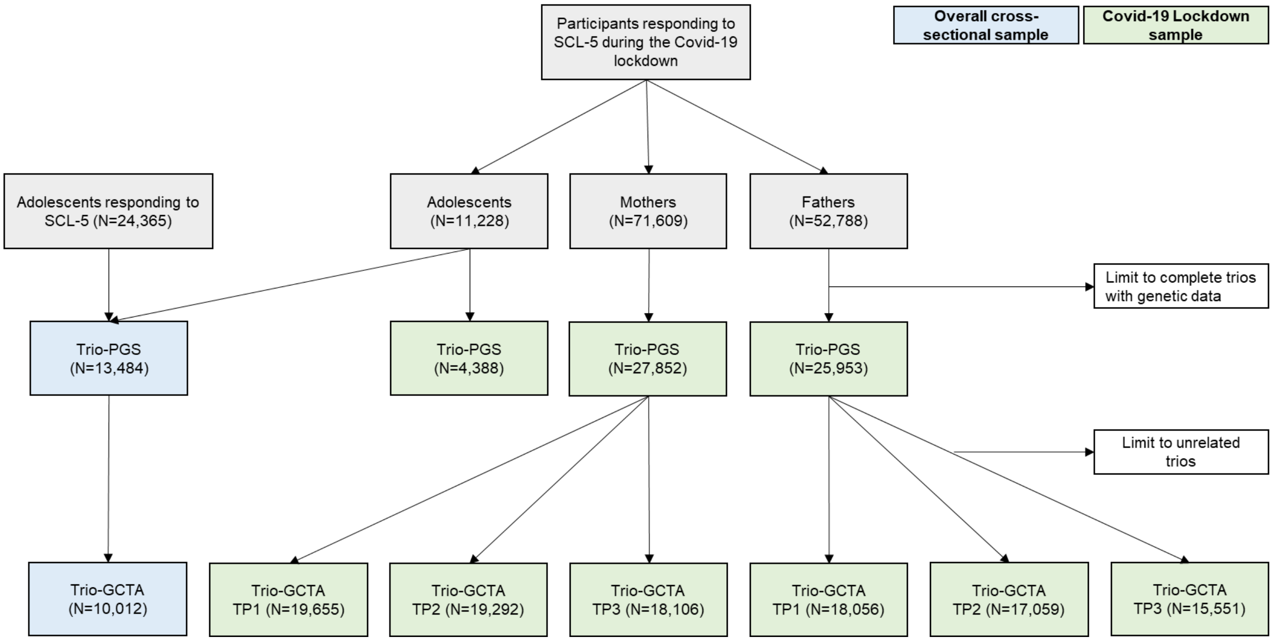

**eMethods.** Further Methodological Details

*Mental distress (SCL-5)*

Due to limited power in the adolescent sample measured during the pandemic, we created an overall mental distress score for all adolescents who had responded to SCL-5 between 14 and 18 years (between 2017-2023) by estimating the mean across available reports (ranging from one to four measurements). All SCL-5 scores were log-transformed and standardized before being included in the analyses. Respondents with fewer than two items answered did not receive an SCL-5 score.

For supplementary PGS-analyses, we included all the available measurements (one to four) for adolescents who had responded to SCL-5 at any time instead of creating a mean score as in the trio-GCTA analyses.

*Household status*

A variable on household status was created based on registered household status from Statistics Norway. Family member trios (mother, father and child) registered in the same household were labeled "living together". Family members not living together were registered as missing. This variable was used to limit the dataset in sensitivity analyses to complete family trios registered as living in the same household.

*Polygenic Scores (PGS)*

PGS for each family member were generated using the LDpred2 software[2], a Bayesian-based method, based on an established pipeline.[3] This pipeline subsets the genetic dataset to HapMap3[4] variants (common SNPs that have been identified across populations) and uses linkage disequilibrium (LD) panels created based on existing LD matrices from UK Biobank. The "LDPred2-auto" option was selected for the PGS scores. To prevent overlap, MoBa participants have been excluded from summary statistics for each trait. PGS were subsequently adjusted for the first 20 genomic principal components, genotype, and imputation batch.

*Statistical analyses - Trio-GCTA*

The variance components estimated in the model can be seen in equation 1.

$$Var(Y) = \sigma^2_m + \sigma^2_p + \sigma^2_o + \sigma_{om} + \sigma_{op} + \sigma^2_e \quad (1)$$

When the adolescent is the focal individual, the $\sigma^2_o$ is interpreted as the direct effect, and the $\sigma^2_m$ and $\sigma^2_p$ are the variance attributed to the maternal and paternal indirect effects, respectively. The $\sigma_{om}$ represents the covariances between the child and maternal indirect effects, and $\sigma_{op}$ are the covariances between the child's direct effect and paternal indirect effects. $\sigma^2_e$ represents the residual variance not explained by the genetic effects. Covariance between maternal and paternal indirect genetic effects $\sigma_{mp}$ is estimated but we do not expect it to contribute to the total variance of the phenotype.

Five models were fun for each focal individual: 1) A full model including all parameters; 2-3) Two models including the direct effect of the focal individual and the indirect effect of each of the other family members (in separate models) in addition the covariance between the direct and indirect effects. For instance, when the adolescent is the focal individual, one model included the child's direct effect, the maternal indirect effect and the covariance between maternal and child effects, and an error term (Model MO), while the second model substituted the maternal with the paternal indirect effect (Model FO); 4) A model with only direct effects (and error term); and 5) a null model including only the error term.

Akaike's Information Criteria (AIC) was used to select the best fitting models. We also reported Bayesian Information Criteria (BIC) and performed Likelihood ratio tests by comparing each model to the full model, estimating whether the nested models provided a significantly worse fit to the data ($\alpha<0.05$). However, due to the use of family data and complex models, BIC and likelihood ratio tests might perform worse in selecting the model closest to the true model[5,6]. We therefore rely on AIC in model selection.

**eTable 2** Variance components included in the six different models. For each focal individual/sample, 5 models were run.

| **Model** | **Variance decomposition** | | |
|---|---|---|---|
| | Child | Mother | Father |
| Full | $\sigma^2_o + \sigma^2_m + \sigma^2_p + \sigma^2_{om} + \sigma^2_{op} + \sigma^2_e$ | $\sigma^2_o + \sigma^2_m + \sigma^2_p + \sigma^2_{om} + \sigma^2_{op} + \sigma^2_e$ | $\sigma^2_o + \sigma^2_m + \sigma^2_p + \sigma^2_{om} + \sigma^2_{op} + \sigma^2_e$ |
| Child and mother (MO) | $\sigma^2_o + \sigma^2_m + \sigma^2_{om} + \sigma^2_e$ | $\sigma^2_o + \sigma^2_m + \sigma^2_{om} + \sigma^2_e$ | |
| Child and father (FO) | $\sigma^2_o + \sigma^2_p + \sigma^2_{op} + \sigma^2_e$ | | $\sigma^2_m + \sigma^2_p + \sigma^2_{mp} + \sigma^2_e$ |
| Mother and father (MF) | | $\sigma^2_m + \sigma^2_p + \sigma^2_{mp} + \sigma^2_e$ | $\sigma^2_m + \sigma^2_p + \sigma^2_{mp} + \sigma^2_e$ |
| Direct | $\sigma^2_o + \sigma^2_e$ | $\sigma^2_o + \sigma^2_e$ | $\sigma^2_o + \sigma^2_e$ |
| Null | $\sigma^2_e$ | $\sigma^2_e$ | $\sigma^2_e$ |

**eTable 3:** Model parameter estimates and model fit statistics for Trio-GCTA model run on mothers' and fathers' mental distress

| Focal individual | | Model | Model parameters (SE) | | | | | | | -2ll | AIC | BIC | Df | p-value |
|---|---|---|---|---|---|---|---|---|---|---|---|---|---|---|
| **Mothers** | | | $\sigma^2_o$ | $\sigma^2_m$ | $\sigma^2_p$ | $\sigma^2_{om}$ | $\sigma^2_{op}$ | $\sigma^2_{mp}$ | $\sigma^2_e$ | | | | | |
| | TP1 | Full | 0.010 (0.022) | 0.091 (0.025) | 0.002 (0.004) | 0.004 (0.020) | 0.003 (0.007) | 0.012 (0.014) | 0.889 (0.025) | 55452.59 | 55538.59 | 55877.71 | 43 | |
| | | MO | 0.010 (0.000) | 0.091 (0.025) | | 0.004 (0.019) | | | 0.889 (0.018) | 55453.76 | 55533.76 | 55849.22 | 40 | 0.760 |
| | | MF | | 0.089 (0.017) | 0.002 (0.000) | | | 0.012 (0.011) | 0.873 (0.020) | 55453.33 | 55533.33 | 55848.79 | 40 | 0.863 |
| | | **Direct** | | **0.099 (0.017)** | | | | | **0.901 (0.017)** | **55455.92** | **55531.92** | **55831.61** | **38** | **0.648** |
| | | Null | | | | | | | 1.0 | 55494.92 | 55568.92 | 55860.72 | 37 | <0.001 |
| | TP2 | Full | 0.001 (0.008) | 0.098 (0.025) | 0.030 (0.024) | -0.006 (0.017) | -0.006 (0.016) | 0.045 (0.018) | 0.882 (0.025) | 54444.99 | 54530.99 | 54869.29 | 43 | |
| | | MO | 0.001 (0.000) | 0.098 (0.021) | | -0.006 (0.010) | | | 0.882 (0.018) | 54454.90 | 54534.90 | 54849.60 | 40 | 0.019 |
| | | **MF** | | **0.095 (0.018)** | **0.029 (0.000)** | | | **0.043 (0.012)** | **0.849 (0.021)** | **54445.21** | **54525.21** | **54839.91** | **40** | **0.973** |
| | | Direct | | 0.095 (0.018) | | | | | 0.905 (0.018) | 54457.61 | 54533.61 | 54832.57 | 38 | 0.027 |
| | | Null | | | | | | | 1.0 | 54490.17 | 54564.17 | 54855.27 | 37 | <0.001 |
| | TP3 | Full | 0.008 (0.036) | 0.094 (0.029) | 0.027 (0.027) | -0.007 (0.026) | 0.000 (0.025) | 0.017 (0.021) | 0.878 (0.031) | 51095.76 | 51181.76 | 51517.33 | 43 | |
| | | MO | 0.008 (0.000) | 0.094 (0.027) | | -0.007 (0.020) | | | 0.878 (0.020) | 51099.09 | 51179.09 | 51491.25 | 40 | 0.343 |
| | | **MF** | | **0.093 (0.019)** | **0.026 (0.000)** | | | **0.017 (0.013)** | **0.865 (0.022)** | **51095.87** | **51175.87** | **51488.03** | **40** | **0.990** |
| | | Direct | | 0.092 (0.019) | | | | | 0.908 (0.019) | 51100.09 | 51176.09 | 51472.65 | 38 | 0.502 |
| | | Null | | | | | | | 1.0 | 51126.80 | 51200.80 | 51489.55 | 37 | <0.001 |
| **Fathers** | | | | | | | | | | | | | | |
| | TP1 | Full | 0.014 (0.014) | 0.011 (0.026) | 0.149 (0.030) | -0.011 (0.015) | -0.045 (0.024) | 0.030 (0.020) | 0.882 (0.027) | 50944.21 | 51030.21 | 51365.66 | 43 | |
| | | FO | 0.014 (0.000) | | 0.149 (0.027) | | -0.045 (0.018) | | 0.882 (0.020) | 50946.55 | 51036.55 | 51338.60 | 40 | 0.051 |
| | | MF | | 0.011 (0.020) | 0.148 (0.012) | | | 0.030 (0.015) | 0.878 (0.023) | 50948.63 | 51028.63 | 51340.68 | 40 | 0.220 |
| | | **Direct** | | | **0.104 (0.020)** | | | | **0.896 (0.020)** | **50948.74** | **51024.74** | **51321.19** | **38** | **0.476** |
| | | Null | | | | | | | 1.0 | 50979.95 | 51053.95 | 51342.60 | 37 | <0.001 |
| | TP2 | Full | 0.016 (0.026) | 0.009 (0.011) | 0.115 (0.032) | 0.003 (0.011) | -0.017 (0.026) | 0.025 (0.019) | 0.873 (0.029) | 48101.90 | 48185.90 | 48511.17 | 42 | |
| | | FO | 0.016 (0.000) | | 0.115 (0.030) | | -0.017 (0.022) | | 0.873 (0.022) | 48105.01 | 48183.01 | 48485.05 | 39 | 0.375 |
| | | MF | | 0.009 (0.020) | 0.112 (0.002) | | | 0.025 (0.015) | 0.853 (0.026) | 48102.86 | 48180.86 | 48482.89 | 39 | 0.812 |
| | | **Direct** | | | **0.102 (0.021)** | | | | **0.898 (0.021)** | **48106.23** | **48180.23** | **48466.77** | **37** | **0.504** |
| | | Null | | | | | | | 1.0 | 48132.36 | 48204.36 | 48483.52 | 36 | <0.001 |
| | TP3 | Full | 0.046 (0.038) | 0.011 (0.013) | 0.101 (0.035) | 0.006 (0.018) | -0.034 (0.031) | 0.024 (0.023) | 0.869 (0.032) | 43918.81 | 44004.81 | 44333.84 | 43 | |
| | | **FO** | **0.046 (0.034)** | | **0.101 (0.001)** | | **-0.034 (0.026)** | | **0.869 (0.024)** | **43921.61** | **44001.61** | **44307.68** | **40** | **0.423** |
| | | MF | | 0.011 (0.022) | 0.099 (0.001) | | | 0.023 (0.016) | 0.849 (0.026) | 43922.15 | 44002.15 | 44308.23 | 40 | 0.341 |
| | | Direct | | | 0.079 (0.022) | | | | 0.921 (0.022) | 43926.03 | 44002.03 | 44292.80 | 38 | 0.205 |
| | | Null | | | | | | | 1.0 | 43939.68 | 44013.68 | 44296.80 | 37 | 0.002 |

**Note:** Bold values indicate the model with lowest Akaike's Information Criteria (AIC) estimate. SE = standard error; -2ll = 2 log likelihood; BIC = Bayesian Information Criteria, df = degrees of freedom.

**eTable 4:** Model parameter estimates and model fit statistics for Trio-GCTA model run on adolescents' mental distress

| Model | Model parameters (SE) | | | | | | | -2ll | AIC | BIC | df | p-value |
|---|---|---|---|---|---|---|---|---|---|---|---|---|
| | $\sigma^2_o$ | $\sigma^2_m$ | $\sigma^2_p$ | $\sigma^2_{om}$ | $\sigma^2_{op}$ | $\sigma^2_{mp}$ | $\sigma^2_e$ | | | | | |
| Full | 0.088 (0.059) | 0.104 (0.049) | 0.017 (0.018) | -0.013 (0.044) | -0.001 (0.027) | 0.041 (0.027) | 0.805 (0.047) | 27055.06 | 27141.06 | 27451.15 | 43 | |
| **MO** | **0.088 (0.002)** | **0.104 (0.049)** | | **-0.013 (0.037)** | | | **0.804 (0.038)** | **27058.63** | **27138.63** | **27427.10** | **40** | **0.311** |
| FO | 0.088 (0.000) | | 0.017 (0.003) | | -0.001 (0.023) | | 0.804 (0.026) | 27064.51 | 27144.51 | 27432.97 | 40 | 0.024 |
| Direct | 0.134 (0.035) | | | | | | 0.866 (0.035) | 27064.66 | 27140.66 | 27414.69 | 38 | 0.087 |
| Null | | | | | | | 1.0 | 27080.52 | 27154.52 | 27421.35 | 37 | <0.001 |

**Note:** Bold values indicate the model with lowest Akaike's Information Criteria (AIC) estimate. SE = standard error; -2ll = 2 log likelihood; BIC = Bayesian Information Criteria, df = degrees of freedom.

**eTable 5:** Standardized beta estimates for PGS on adolescents', mothers' and fathers' mental distress

| | | Anxiety | | Depression | | ADHD | | Neuroticism | | Anorexia | |
|---|---|---|---|---|---|---|---|---|---|---|---|
| | | Std. estimate (SE) | P-value | Std. estimate (SE) | P-value | Std. estimate (SE) | P-value | Std. estimate (SE) | P-value | Std. estimate (SE) | P-value |
| **Adolescents** | Child PGS (direct) | 0.025 (0.019) | 0.190 | 0.084 (0.019) | **<0.001*** | 0.032 (0.019) | 0.090 | 0.072 (0.019) | **<0.001*** | -0.034 (0.019) | 0.079 |
| | Mother PGS (indirect) | 0.005 (0.016) | 0.740 | 0.026 (0.017) | 0.120 | 0.034 (0.017) | **0.037** | -0.009 (0.016) | 0.586 | 0.013 (0.017) | 0.456 |
| | Father PGS (indirect) | 0.016 (0.016) | 0.332 | 0.029 (0.016) | 0.075 | 0.021 (0.017) | 0.200 | 0.031 (0.016) | 0.054 | 0.025 (0.017) | 0.139 |
| | Time | -0.004 (0.0005) | **<0.001*** | -0.004 (0.0005) | **<0.001*** | -0.004 (0.0005) | **<0.001*** | -0.004 (0.0005) | **<0.001*** | -0.004 (0.0005) | **<0.001*** |
| | Child PGS*time | 0.00001 (0.0007) | 0.986 | -0.001 (0.0007) | 0.166 | 0.001 (0.0007) | 0.109 | -0.0005 (0.0007) | 0.447 | -0.0002 (0.0007) | 0.773 |
| | Mother PGS*time | -0.0001 (0.0006) | 0.860 | 0.001 (0.0006) | 0.104 | 0.0005 (0.0006) | 0.409 | -0.0001 (0.0006) | 0.866 | 0.0002 (0.0006) | 0.735 |
| | Father PGS*time | -0.0003 (0.0006) | 0.572 | 0.0008 (0.0006) | 0.176 | -0.0009 (0.0006) | 0.149 | 0.0001 (0.0006) | 0.799 | 0.0005 (0.0006) | 0.416 |
| **Mothers** | Mother PGS (direct) | 0.048 (0.007) | **<0.001*** | 0.108 (0.007) | **<0.001*** | 0.027 (0.007) | **<0.001*** | 0.105 (0.007) | **<0.001*** | 0.030 (0.007) | **<0.001*** |
| | Father PGS (indirect) | -0.002 (0.007) | 0.802 | 0.014 (0.007) | **0.042** | 0.004 (0.007) | 0.529 | 0.010 (0.007) | 0.129 | -0.005 (0.007) | 0.480 |
| | Child PGS (indirect) | 0.017 (0.008) | **0.030** | -0.003 (0.008) | 0.745 | 0.008 (0.008) | 0.314 | -0.002 (0.008) | 0.838 | -0.001 (0.008) | 0.890 |
| | Time | -0.007 (0.0002) | **<0.001*** | -0.007 (0.0002) | **<0.001*** | -0.007 (0.0002) | **<0.001*** | -0.007 (0.0002) | **<0.001*** | -0.007 (0.0002) | **<0.001*** |
| | Mother PGS*time | -0.0001 (0.0002) | 0.727 | 0.0001 (0.0002) | 0.681 | 0.0002 (0.0002) | 0.355 | 0.0004 (0.0002) | **0.039** | 0.00001 (0.0002) | 0.976 |
| | Father PGS*time | 0.00004 (0.0002) | 0.848 | 0.0003 (0.0002) | 0.141 | 0.0002 (0.0002) | 0.226 | 0.0001 (0.0002) | 0.498 | -0.0004 (0.0002) | 0.077 |
| | Child PGS*time | 0.00003 (0.0002) | 0.888 | -0.00003 (0.0002) | 0.893 | 0.0002 (0.0002) | 0.286 | -0.0003 (0.0002) | 0.223 | 0.0005 (0.0002) | 0.052 |
| **Fathers** | Father PGS (direct) | 0.049 (0.007) | **<0.001*** | 0.103 (0.007) | **<0.001*** | 0.037 (0.007) | **<0.001*** | 0.098 (0.007) | **<0.001*** | 0.025 (0.007) | **<0.001*** |
| | Mother PGS (indirect) | 0.0002 (0.007) | 0.977 | 0.023 (0.007) | **0.001*** | 0.029 (0.007) | **<0.001*** | 0.013 (0.007) | 0.060 | 0.006 (0.007) | 0.411 |
| | Child PGS (indirect) | 0.010 (0.008) | 0.231 | -0.005 (0.008) | 0.565 | -0.020 (0.008) | **0.016** | -0.003 (0.008) | 0.743 | -0.006 (0.008) | 0.464 |
| | Time | -0.004 (0.0002) | **<0.001*** | -0.004 (0.0002) | **<0.001*** | -0.004 (0.0002) | **<0.001*** | -0.004 (0.0002) | **<0.001*** | -0.004 (0.0002) | **<0.001*** |
| | Father PGS*time | 0.00001 (0.0002) | 0.948 | 0.0001 (0.0002) | 0.751 | 0.0004 (0.0002) | 0.090 | -0.0001 (0.0002) | 0.776 | 0.0001 (0.0002) | 0.521 |
| | Mother PGS*time | -0.0001 (0.0002) | 0.489 | -0.0001 (0.0002) | 0.640 | -0.0001 (0.0002) | 0.566 | 0.0002 (0.0002) | 0.460 | 0.0002 (0.0002) | 0.344 |
| | Child PGS*time | -0.00007 (0.0002) | 0.768 | 0.0002 (0.0002) | 0.458 | -0.0001 (0.0002) | 0.777 | -0.0002 (0.0002) | 0.377 | -0.0002 (0.0002) | 0.412 |

**Note:** Bold indicate p-values less than 0.05 and * indicate significant p-values after FDR correction. SE = standard error.

**eTable 6:** Supplementary analyses – Standardized beta estimates for PGS on adolescents' mental distress

| | Anxiety | | Depression | | ADHD | | Neuroticism | | Anorexia | |
|---|---|---|---|---|---|---|---|---|---|---|
| | Std. estimate (SE) | P-value | Std. estimate (SE) | P-value | Std. estimate (SE) | P-value | Std. estimate (SE) | P-value | Std. estimate (SE) | P-value |
| Direct – child | 0.033 (0.011) | **0.003*** | 0.099 (0.011) | **<0.001*** | 0.044 (0.011) | **<0.001*** | 0.085 (0.011) | **<0.001*** | 0.004 (0.011) | 0.772 |
| Indirect – maternal | 0.008 (0.010) | 0.427 | 0.021 (0.010) | **0.033** | 0.018 (0.010) | 0.057 | -0.014 (0.010) | 0.148 | 0.001 (0.010) | 0.893 |
| Indirect – paternal | 0.009 (0.010) | 0.339 | 0.016 (0.010) | 0.092 | -0.004 (0.010) | 0.715 | 0.009 (0.010) | 0.379 | 0.012 (0.010) | 0.210 |

**Note:** Bold indicate p-values less than 0.05 and * indicate significant p-values after FDR correction. SE = standard error.

**eFigure 2** Standardized beta estimates for PGS on adolescents' mental distress.

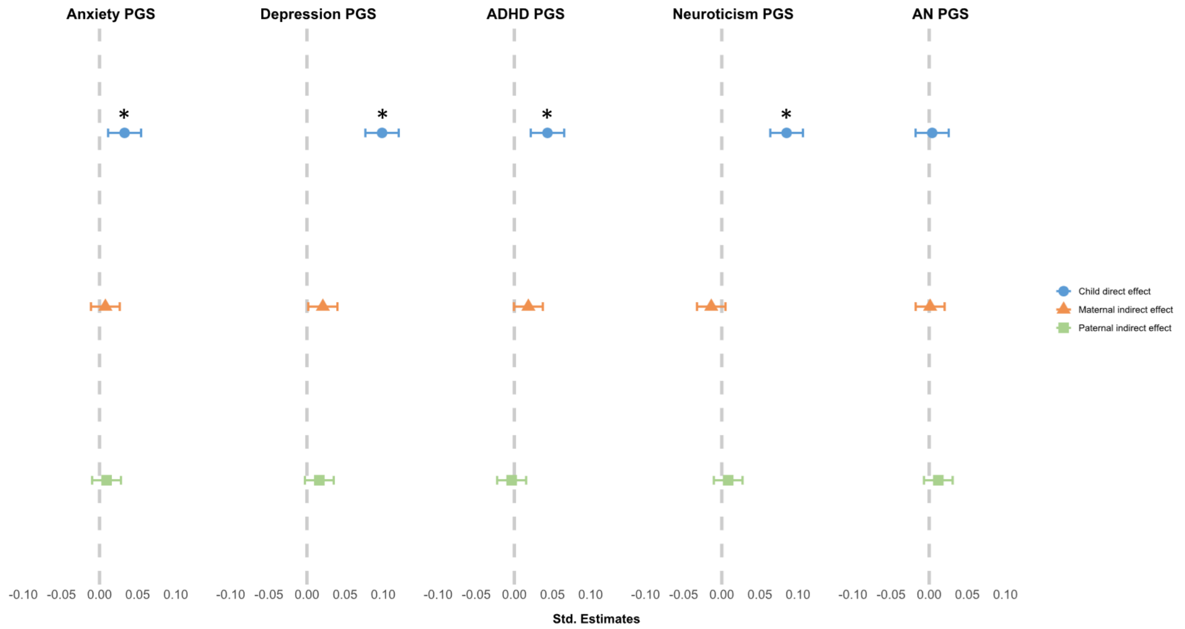

**Note:** Blue circles show direct genetic effects , and indirect effects are indicated by orange triangles (maternal) and green squares (paternal). * indicates significant p-values after FDR correction.

**eTable 7:** Sensitivity analyses - Standardized beta estimates for PGS on adolescents', mothers' and fathers' mental distress in trios restricted to living in the same household

| | | Anxiety | | Depression | | ADHD | | Neuroticism | | Anorexia | |
|---|---|---|---|---|---|---|---|---|---|---|---|
| | | Std. estimate (SE) | P-value | Std. estimate (SE) | P-value | Std. estimate (SE) | P-value | Std. estimate (SE) | P-value | Std. estimate (SE) | P-value |
| **Adolescents** | Child PGS (direct) | 0.020 (0.022) | 0.349 | 0.081 (0.022) | **<0.001*** | 0.046 (0.021) | **0.031** | 0.046 (0.021) | **0.031** | -0.027 (0.022) | 0.208 |
| | Mother PGS (indirect) | -0.009 (0.018) | 0.629 | 0.027 (0.019) | 0.152 | 0.032 (0.019) | 0.090 | -0.032 (0.019) | 0.586 | 0.021 (0.019) | 0.261 |
| | Father PGS (indirect) | 0.007 (0.018) | 0.716 | 0.018 (0.019) | 0.342 | 0.006 (0.019) | 0.750 | 0.031 (0.019) | 0.054 | 0.014 (0.019) | 0.445 |
| | Time | -0.004 (0.0004) | **<0.001*** | -0.005 (0.0004) | **<0.001*** | -0.005 (0.0004) | **<0.001*** | -0.005 (0.0004) | **<0.001*** | -0.005 (0.0004) | **<0.001*** |
| | Child PGS*time | 0.0001 (0.0006) | 0.092 | -0.001 (0.0006) | 0.118 | 0.0002 (0.0006) | 0.788 | 0.0002 (0.0006) | 0.788 | 0.0006 (0.0006) | 0.333 |
| | Mother PGS*time | -0.0003 (0.0005) | 0.522 | 0.001 (0.0005) | **0.026** | 0.0007 (0.0005) | 0.184 | 0.0007 (0.0005) | 0.184 | -0.0005 (0.0005) | 0.369 |
| | Father PGS*time | -0.0002 (0.0005) | 0.772 | 0.0004 (0.0005) | 0.460 | -0.0003 (0.0005) | 0.566 | -0.0003 (0.0005) | 0.566 | -0.0004 (0.0005) | 0.486 |
| **Mothers** | Mother PGS (direct) | 0.049 (0.007) | **<0.001*** | 0.107 (0.008) | **<0.001*** | 0.023 (0.008) | **0.004*** | 0.100 (0.008) | **<0.001*** | 0.027 (0.008) | **<0.001*** |
| | Father PGS (indirect) | 0.001 (0.007) | 0.912 | 0.019 (0.008) | **0.013** | -0.002 (0.008) | 0.798 | 0.015 (0.008) | 0.129 | -0.008 (0.008) | 0.334 |
| | Child PGS (indirect) | 0.013 (0.008) | 0.138 | -0.006 (0.009) | 0.510 | 0.009 (0.009) | 0.332 | -0.002 (0.009) | 0.838 | 0.004 (0.009) | 0.653 |
| | Time | -0.007 (0.0002) | **<0.001*** | -0.007 (0.0002) | **<0.001*** | -0.007 (0.0002) | **<0.001*** | -0.007 (0.0002) | **<0.001*** | -0.007 (0.0002) | **<0.001*** |
| | Mother PGS*time | -0.0001 (0.0002) | 0.684 | -0.0003 (0.0002) | 0.295 | -0.000001 (0.0002) | 0.995 | 0.0004 (0.0002) | 0.124 | -0.00004 (0.0002) | 0.687 |
| | Father PGS*time | 0.00006(0.0002) | 0.813 | 0.0002 (0.0002) | 0.390 | 0.0003 (0.0002) | 0.164 | 0.0001 (0.0002) | 0.663 | -0.0004 (0.0002) | 0.065 |
| | Child PGS*time | -0.0001 (0.0002) | 0.661 | 0.0002 (0.0003) | 0.395 | 0.0004 (0.0003) | 0.119 | -0.0004 (0.0003) | 0.124 | 0.0006 (0.0003) | **0.032** |
| **Fathers** | Father PGS (direct) | 0.054 (0.008) | **<0.001*** | 0.101 (0.007) | **<0.001*** | 0.029 (0.008) | **<0.001*** | 0.102 (0.008) | **<0.001*** | 0.019 (0.007) | **0.016*** |
| | Mother PGS (indirect) | 0.002 (0.008) | 0.834 | 0.023 (0.007) | **0.006*** | 0.030 (0.008) | **<0.001*** | 0.018 (0.008) | **0.030** | 0.006 (0.007) | 0.442 |
| | Child PGS (indirect) | 0.002 (0.009) | 0.853 | -0.005 (0.008) | 0.727 | -0.015 (0.009) | 0.105 | -0.011 (0.009) | 0.237 | -0.0004 (0.008) | 0.965 |
| | Time | -0.004 (0.0002) | **<0.001*** | -0.005 (0.0001) | **<0.001*** | -0.005 (0.0002) | **<0.001*** | -0.005 (0.0002) | **<0.001*** | -0.005 (0.0002) | **<0.001*** |
| | Father PGS*time | -0.00005 (0.0002) | 0.847 | -0.0001 (0.0002) | 0.770 | 0.0004 (0.0002) | 0.106 | 0.0002 (0.0002) | 0.513 | 0.0001 (0.0002) | 0.560 |
| | Mother PGS*time | -0.0002 (0.0002) | 0.434 | -0.0001 (0.0002) | 0.557 | 0.0001 (0.0002) | 0.850 | 0.0002 (0.0002) | 0.504 | 0.0003 (0.0002) | 0.182 |
| | Child PGS*time | 0.0001 (0.0003) | 0.770 | 0.0001 (0.0003) | 0.759 | -0.0002 (0.0003) | 0.482 | -0.0002 (0.0003) | 0.518 | -0.0004 (0.0003) | 0.197 |

**Note:** Bold indicate p-values less than 0.05 and * indicate significant p-values after FDR correction. SE = standard error.

### eReferences

1. Willroth EC, Atherton OE. Best laid plans: A guide to reporting preregistration deviations. *Advances in Methods and Practices in Psychological Science*. 2024;7(1):25152459231213802.
2. Privé F, Arbel J, Vilhjálmsson BJ. LDpred2: better, faster, stronger. *Bioinformatics*. 2020;36(22-23):5424-5431.
3. Allegrini A. *AndreaAllegrini/LDpred2[R]*. https://github.com/AndreAllegrini/LDpred2
4. Consortium IH. Integrating common and rare genetic variation in diverse human populations. *Nature*. 2010;467(7311):52.
5. Vrieze SI. Model selection and psychological theory: a discussion of the differences between the Akaike information criterion (AIC) and the Bayesian information criterion (BIC). *Psychol Methods*. 2012;17(2):228.
6. Dominicus A, Skrondal A, Gjessing HK, Pedersen NL, Palmgren J. Likelihood ratio tests in behavioral genetics: problems and solutions. *Behav Genet*. 2006;36:331-340.
